# Magnitude of perinatal mortality in sub-Saharan Africa since 2000: a systematic review and meta-analysis protocol

**DOI:** 10.1101/2025.09.22.25336421

**Authors:** Musonda Makasa, Patrick Kaonga, Choolwe Jacobs, Mpundu Makasa, Patrick Musonda, Joseph Mumba Zulu, Mercy Wamunyima Monde, Bellingtone Vwalika

## Abstract

**Background:** A lot of unborn and newly born babies continue to die during pregnancy, childbirth and in the first week of life. Over 5,1 million stillbirths and neonatal deaths occur globally each year with 98% of these in low-middle-income countries. Since the Millennium Development Goals (MDG) era, global perinatal mortality reduced from 5.7 million in 2000 to 4.1 million by 2015. The global stillbirth rate which averaged 24.7 in 2000 declined to 18.4 per 1000 live births by 2015. Neonatal mortality rate equally showed a downward trend from 37 in 1990 to 19 deaths per 1000 live births by 2016. In spite of this, sub-Saharan Africa (SSA) has continued to record the highest rate as 2% of the 45% data collected is in high income countries. The average perinatal mortality rate is at 30.3 per 1000 live births with Ethiopia reportedly highest at 49. The main aim of this study is to determine the magnitude of perinatal mortality in SSA since 2000.

**Methods:** The databases searched are EMBASE, Scopus, and CINAHIL the followed by grey literature sources including ProQuest Global Thesis, and Dissertation, and Google Scholar. The articles for the publications made from January 2000 to December, 2023. Quality of the articles check was with Joanna Briggs Institute quality assessment tool. Three reviewers screened all retrieved articles, extracted, and then critically appraised all identified studies. Stata software version 17 will be used.

**Discussion:** The review will provide a detailed summary of the magnitude of perinatal mortality in SSA since 2000. The review will synthesis available observational studies on perinatal mortality to identify pooled estimates.

**Systematic review registration:** PROSPERO CRD42023437432

## Background

Perinatal mortality remains a major global health challenge, reflecting both the burden of stillbirths and early neonatal deaths, with the vast majority of these preventable losses occurring in low- and middle-income countries (LMICs). Globally, approximately 5.1 million stillbirths and neonatal deaths occur each year, and 98% of these take place in LMICs [1]. Recent global estimates indicate that more than 50 million stillbirths have occurred since the year 2000, including 1.9 million in 2021 alone. Alongside neonatal mortality, stillbirths remain key indicators of perinatal health, and each preventable loss represents a setback in achieving the 2030 Agenda for Sustainable Development and upholding the Convention on the Rights of the Child, particularly its fundamental principle of the right to survival [2].

Despite global commitments, efforts to prevent perinatal mortality have achieved limited success. Stillbirths, in particular, remain underrepresented in global data tracking, social recognition, and programmatic action, which has contributed to slow progress. Perinatal mortality is defined by the World Health Organization (WHO) as the loss of a foetus after 28 weeks of gestation, birthweight ≥1000 g, or death of a neonate within the first seven days of life [3]. It is closely linked to maternal health; for every maternal death, an estimated ten perinatal deaths occur, with two-thirds of maternal death causes overlapping with those of perinatal mortality [4].

Global initiatives have sought to address this challenge. The Millennium Development Goals (MDGs) targeted child and maternal health, and the subsequent Sustainable Development Goals (SDGs) committed to reducing neonatal mortality to at least 12 deaths per 1,000 live births by 2030 [5]. Progress has been made: global perinatal mortality fell from 5.7 million in 2000 to 4.1 million in 2015, and the stillbirth rate declined from 24.7 to 18.4 per 1,000 live births over the same period [6, 7]. Similarly, global neonatal mortality significantly declined from approximately 36.6 deaths per 1000 live births in 1990 to around 18 per 1,000 live in 2017, representing a 51% reduction over this period [8]. However, nearly half of the available data originate from high-income countries, which contribute less than 2% of the global burden [9]. Sub-Saharan Africa (SSA) bears the highest perinatal mortality rates globally. For example, Nigeria has been reported to have a rate of 40.9 per 1,000 live births, the highest in West Africa, while Ethiopia reports nearly 49 per 1,000 [10, 11]. A recent systematic review estimated the pooled perinatal mortality rate in SSA at 58.35 per 1,000 live births (observed) and 42.95 per 1,000 live births (adjusted), with southern Africa reporting lower rates at around 30.3 [12].

According to WHO, perinatal mortality reflects not only the socioeconomic status of a country but also the availability, accessibility, and quality of maternal, obstetric, and neonatal services [3, 13]. Despite global and regional initiatives, the burden in SSA remains unacceptably high, and reductions have been slow compared to other regions. In light of this, the proposed systematic review and meta-analysis will synthesize available evidence to estimate the pooled prevalence of perinatal mortality and its predictors in SSA from 2000 onwards, coinciding with the global commitment to the MDGs. This will generate updated evidence to inform policy, guide programming, and strengthen strategies aimed at reducing preventable perinatal deaths in the region.

### Study Aim

To estimate the pooled prevalence of perinatal mortality and the pooled effect sizes of its predictors in SSA, with 95% confidence intervals, using random-effects meta-analysis.

### Specific Objectives

1. To estimate the pooled prevalence of perinatal mortality in SSA, with 95% confidence intervals, using random-effects meta-analysis.
2. To determine the pooled effect sizes of predictors of perinatal mortality in SSA, expressed as odds ratios or risk ratios with 95% confidence intervals, using random-effects models.

## Methods

This systematic review and meta-analysis will be conducted in accordance with the Preferred Reporting Items for Systematic Reviews and Meta-Analyses (PRISMA) 2020 guidelines [14]. The protocol has been registered in the PROSPERO database (CRD42023437432).

### Search Strategy

A systematic literature search will be conducted in PubMed and iteratively adapted for other databases, including EMBASE, Scopus, and CINAHL. Grey literature sources will also be searched, including ProQuest Global Theses and Dissertations, WHO IRIS, and Google Scholar. The search will cover publications between January 2000, and December 2023. To ensure completeness, reference lists of included studies will also be screened. The search strategy will combine Medical Subject Headings (MeSH) and free-text terms using Boolean operators (“AND,” “OR”), truncation, and synonyms to balance precision and sensitivity. Core search terms will include: *perinatal mortality, perinatal death, stillbirth, intrauterine fetal/foetal death, early neonatal death, early neonatal mortality, incidence, prevalence, ratio, risk factors, rate, magnitude*, and *sub-Saharan Africa (Africa south of the Sahara, SSA)*.

### Study Identification and Selection

All retrieved citations will be imported into EndNote for deduplication and then screened in Rayyan.ai. Four reviewers (MM, KC, JS, and one additional reviewer) will independently screen titles and abstracts, followed by full-text screening by three reviewers (MM, KS, MK). Disagreements will be resolved by consensus, with arbitration by a third reviewer if necessary. Reasons for exclusion at the full-text stage will be documented, and the overall selection process will be summarized in a PRISMA flow diagram.

### Eligibility Criteria

This review will include observational analytical study designs, specifically cohort, case–control, and cross-sectional studies, that report on perinatal mortality—defined as stillbirth and/or early neonatal mortality—or its predictors in SSA. To align with the era of global commitments under the MDGs and SDGs, only studies published between January 2000, and December 2023, will be considered. Eligible studies must provide sufficient data to calculate either prevalence estimates or effect sizes for the outcomes of interest.

Studies will be excluded if they are qualitative in nature, systematic or narrative reviews, case reports, or conference abstracts. We will also exclude studies not in English language and that do not provide extractable data on perinatal mortality or its predictors, sample size justification as well as not conducted outside sub-Saharan Africa.

### Selection of studies

Figure 1 hereafter presents the planned flow diagram outlining the search and study selection procedure. The process begins with the systematic identification of records from multiple electronic databases and additional sources. After removal of duplicates, titles and abstracts will be screened against predefined eligibility criteria. Studies deemed potentially relevant will undergo full-text review to assess inclusion. Finally, the number of studies meeting all criteria will be documented and included in the systematic review and meta-analysis. This stepwise approach follows the PRISMA 2020 guidelines to ensure transparency and reproducibility in the review process.

### PRISMA Flowchart

**Figure 1.**
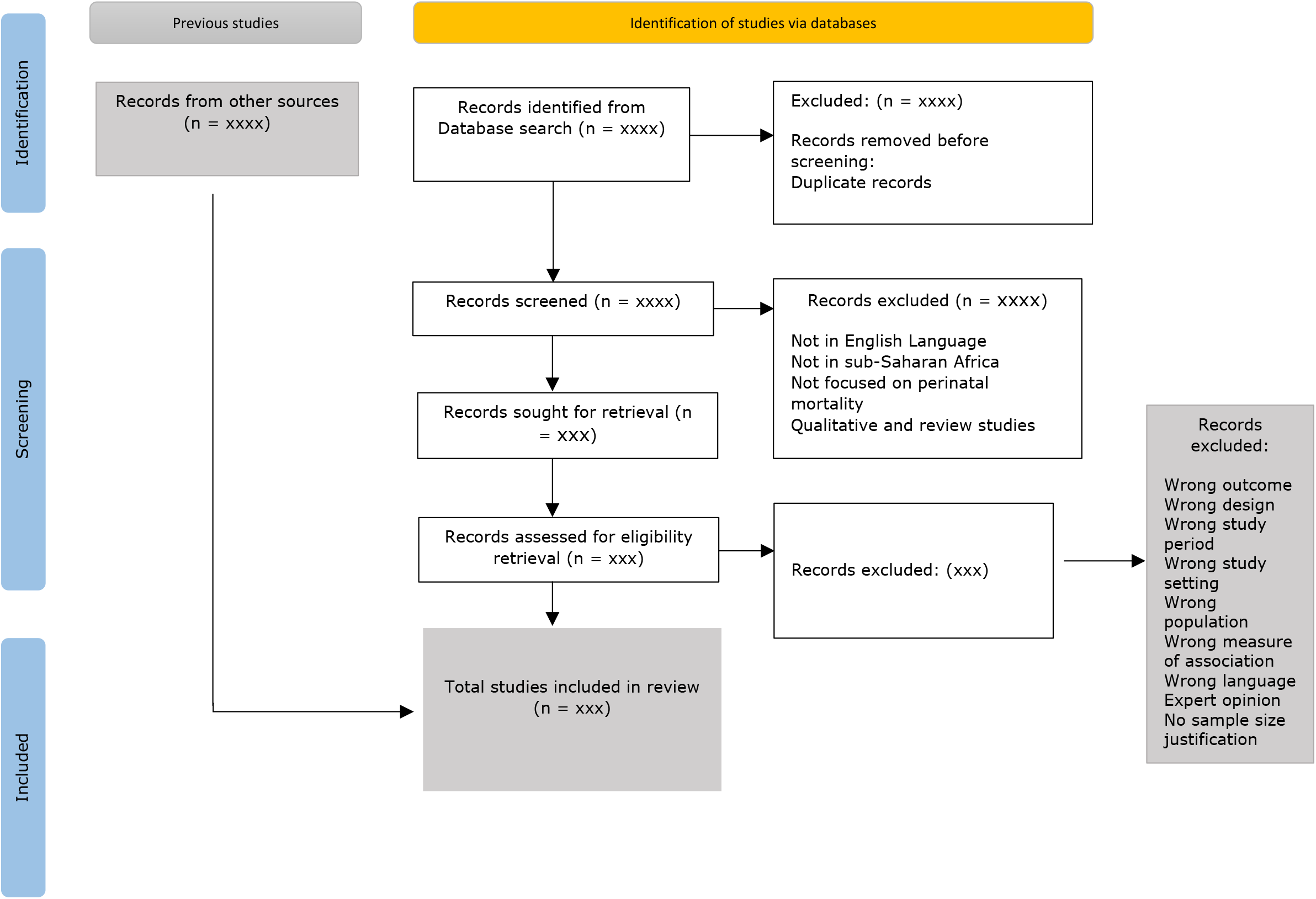
The figure illustrates the flow diagram of search and study selection procedure to be applied.

### Data Extraction and Management

Data will be extracted using a piloted Excel form by three independent reviewers. Extracted information will include study characteristics (author, year, country, region, design, setting, sample size), prevalence of perinatal mortality, predictors assessed, effect estimates (odds ratios, risk ratios, or prevalence), and confidence intervals. Discrepancies will be resolved by consensus, with input from a statistician if needed.

### Risk of Bias Assessment

The methodological quality of included studies will be assessed using the Joanna Briggs Institute (JBI) critical appraisal tools appropriate for each study design [15]. Assessment criteria will include representativeness of the population, appropriateness of study design, adequacy of sample size, outcome measurement, and statistical analysis. Only studies judged to be of at least moderate quality will be included in the meta-analysis.

### Data Synthesis and Statistical Analysis

A narrative summary of study characteristics will be provided, supported by tables and figures. Meta-analysis will be performed in Stata version 17. For prevalence estimates, data will be transformed using the Freeman–Tukey double arcsine method to stabilize variance. For predictors, odds ratios and risk ratios will be log-transformed prior to pooling.

Pooled prevalence and pooled effect sizes (odds ratios or risk ratios) with 95% confidence intervals will be calculated using a random-effects model (DerSimonian–Laird). Between-study heterogeneity will be quantified using the I^2^ statistic and τ^2^. Thresholds of 25%, 50%, and 75% will be interpreted as low, moderate, and high heterogeneity, respectively. Publication bias will be assessed using funnel plots and Egger’s test, with p < 0.10 considered significant. If evidence of bias is detected, the Duval and Tweedie trim-and-fill method will be applied.

### Subgroup and Sensitivity Analyses

Subgroup analyses will be conducted by study region, year of publication, study setting, and study quality. Sensitivity analyses will be performed to test the robustness of pooled estimates by excluding outlier studies and comparing results with and without them.

## Data Availability

No primary data are associated with this protocol. The planned systematic review and meta-analysis will use data extracted from previously published studies and publicly available sources. Once the review is completed, all extracted data supporting the findings will be deposited in the University of Zambia Library repository and made fully accessible. The PROSPERO registration record (CRD42023437432) and PRISMA-P checklist are provided as supporting information for this protocol.

## Ethics and Dissemination

This review will synthesize data from published studies and publicly available sources. No primary data collection will be undertaken, and therefore ethical approval is not required. The protocol is registered in PROSPERO (CRD42023437432). Findings will be disseminated through publication in a peer-reviewed journal and presentations at national and international conferences.

## Declarations

### Consent for publication

Not applicable as the protocol does not contain any person’s individual’s data in any form.

### Authors’ information

^1^University of Zambia, School of Public Health in the Department of Epidemiology and Biostatistics.

^2^Honorary lecturer of Obstetrics and Gynaecology at the University of Zambia, School of Medicine, Department of Obstetrics and Gynaecology, Lusaka, Zambia.

### Authors’ contributions

MM: Conceptualisation of the study, literature review, and manuscript writing. PK supervision, conceptualisation and methodology. CJ: supervision and contextualisation MM: study conceptualisation. JMZ: Study conceptualisation. PM: Methodology MWM: data search. BV: overall supervision, conceptualisation, and content analysis. All authors read and approved the final version of the manuscript.

## Acknowledgements

Acknowledgement and immense appreciation goes to all the authors’ for the time and contributions made towards this protocol development. Secondly, credit and appreciation should also be accorded to the University of Zambia Schools of Public Health and Medicine (Department of Obstetrics and Gynaecology), and the University Teaching Hospital – Women and Newborn Hospital for providing the platform to make this process possible.

